# Automatic segmentation of choroid plexus using deep learning across neurodegenerative diagnoses in the multi-site COMPASS-ND Study

**DOI:** 10.64898/2026.05.14.26353194

**Authors:** Manpreet Singh, Fanta Dabo, Lianne J. Trigiani, David Araujo, Sridar Narayanan, AmanPreet Badhwar

## Abstract

The choroid plexus (ChP) plays a central role in cerebrospinal fluid production, immune signaling, and metabolic clearance, and has emerged as a potential imaging biomarker of neurodegeneration. However, accurate and scalable quantification of ChP volume remains challenging due to its complex morphology and low contrast on conventional MRI. The Automatic Segmentation of Choroid Plexus (ASCHOPLEX), a deep learning framework originally trained on healthy controls and multiple sclerosis cohorts, has not been systematically evaluated in neurodegenerative populations.

Using T1-weighted MRI from the multi-center COMPASS-ND study, we assessed standard ASCHOPLEX performance in cognitively unimpaired (CU), Alzheimer’s disease (AD), and Parkinson’s disease (PD) participants (N = 30), followed by fine-tuning using expert manual segmentations (N = 60). Segmentation accuracy was evaluated using Dice, Jaccard, precision, and recall. The fine-tuned model was then applied to a larger cohort (N = 277) to derive normalized ChP volumes, which were compared across diagnostic groups using linear regression models.

Fine-tuning significantly improved segmentation accuracy across all metrics (Dice: 0.45 to 0.84; Jaccard: 0.32 to 0.73; all *p* < 0.0001), enabling robust ChP delineation across sites and conditions. In the full cohort, normalized ChP volume was significantly higher in AD compared with CU and PD (*p* < 0.0001), while PD did not differ from CU (*p* = 0.31).

These findings demonstrate that dataset-specific adaptation is essential for deploying deep learning segmentation models in heterogeneous neuroimaging cohorts. The refined ASCHOPLEX framework enables scalable ChP quantification and supports its use as a structural imaging marker in neurodegenerative disease.

## 1. Introduction

There is growing interest in the choroid plexus (ChP) and its volume as an early imaging marker of neurodegenerative disease^1–3^. Situated within the ventricular system, the ChP is composed of a monolayer of epithelial cells that forms the blood-cerebrospinal fluid barrier (BCSFB)^4^. Beyond being the primary producer of CSF, it regulates brain homeostasis by supporting nutrient transport, controlling molecular trafficking through protein secretion, and facilitating glymphatic and metabolic clearance pathways^4–6^. Disruptions in these processes - particularly BCSFB dysfunction and impaired clearance - have been increasingly linked to neurodegenerative conditions^7–9^, including Alzheimer’s and Parkinson’s disease, where structural alterations such as ChP enlargement have been reported^1,2,7,10–13^. Accordingly, ChP volume has emerged as a promising early imaging marker of neurodegenerative disease, potentially capturing structural changes that precede detectable alterations in surrounding brain tissue^14,15^.

Accurate and scalable quantification of ChP volume remains challenging due to its heterogeneous relaxometry characteristics^16^. T1-weighted (T1w) MRI is the standard non-invasive method for ChP imaging and segmentation in large cohorts, offering advantages over contrast-based approaches (*e*.*g*., gadolinium-enhanced MRI), which require exogenous contrast agents and are less suitable for repeated measurements^17^. Manual segmentation, while considered the gold standard, is labor-intensive and prone to inter-rater variability due to the ChP’s complex morphology^1^. Automated approaches, including FreeSurfer^1,18^ and Gaussian Mixture Models^1,18^, offer faster alternatives but often mislabel tissue or underestimate boundaries^1,17,19^; issues exacerbated by atrophy, ventricular enlargement, and variability in imaging protocols.

Recent advances in deep learning have enabled more reliable neuroanatomical segmentation across diverse MRI acquisition settings, including differences in scanner type, imaging sequence, and protocol^20,21^. Frameworks such as nnU-Net^22^, and UNETR^23^ leverage self-configuring architectures^22^ and transformer-based attention mechanisms^23^ to advance medical image segmentation. However, their application to small, low-contrast structures like the ChP remains limited and typically requires dataset-specific retraining^19,23^. The Automatic Segmentation of Choroid Plexus (ASCHOPLEX)^19^ model combines an ensemble of nnU-Net and UNETR architectures with optimized loss functions, achieving superior accuracy, robustness, and adaptability through a fine-tuning module^19^. Originally trained on datasets comprising healthy controls and patients with multiple sclerosis, its performance in neurodegenerative populations has not been evaluated.

In this study, we evaluate the performance of ASCHOPLEX across cognitively unimpaired (CU), AD, and PD participants of the pan-Canadian Comprehensive Assessment of Neurodegeneration and Dementia (COMPASS-ND) cohort to assess its real-world generalizability and robustness across distinct neurodegenerative conditions. Through fine-tuning, we further refine the model to account for diagnostic and MRI protocol heterogeneity.

Using the fine-tuned model, we quantify ChP volumes from T1-w images and compare normalized ChP volumes across diagnostic categories. By establishing this validated framework, our work supports scalable and reliable ChP quantification in large neurodegenerative cohorts and provides a tool for investigating ChP structural alterations as a potential imaging marker of neurodegeneration.

## 2. Methods

### 2.1 Study cohort and data selection

Data were sourced from the Canadian Consortium on Neurodegeneration in Aging (CCNA) COMPASS-ND study^24^, a national multi-center initiative involving 32 imaging sites across Canada. This study includes a total of 1,173 participants spread across different diagnostic categories, including CU, AD (with varying levels of vascular brain injury), and PD. All participants provided written informed consent, and ethics approval was obtained at each recruiting center^24^.

For ASCHOPLEX training and validation, 60 participants (20 CU, 20 AD, 20 PD) were pseudo-randomly selected from COMPASS-ND to ensure equal representation across diagnostic categories for age and sex (Table 1). Testing was done on a separate sample of 30 participants (10 CU, 10 AD, 10 PD). To maximize generalizability testing, participants with atypical ventricular morphology or known segmentation operational challenges were retained in the test set. The remaining COMPASS-ND participants classified as CU, AD or PD (70 CU, 109 AD, 98 PD, Table 2) were subsequently processed using the fine-tuned ASCHOPLEX model to obtain automated ChP segmentations for volumetric analyses.

**Table 1:** Demographic characteristics of the COMPASS-ND study cohort used for fine tuning and testing of ChP segmentation. Participants are shown by diagnostic category, with total sample size and allocation to training/validation and testing sets, mean age ± standard deviation (SD), and sex distribution (female/male). *Abbreviations:* AD : Alzheimer’s disease; ChP: choroid plexus; CU : cognitively unimpaired; PD : Parkinson’s disease.

| Diagnostic category | $N^{\text{Total}}$ ($N^{\text{Training and validation}}$, $N^{\text{testing}}$) | Age (mean ± SD) | Sex (F/M) |
| --- | --- | --- | --- |
| CU | 30 (20,10) | 68.4 ± 5.3 | 15 / 15 |
| AD | 30 (20,10) | 72.4 ± 6.0 | 16 / 14 |
| PD | 30 (20,10) | 67.5 ± 7.9 | 15 / 15 |
| **Total cohort** | 90 (60,30) | 69.4 ± 6.8 | 46 / 44 |

**Table 2:** Demographic characteristics of the COMPASS-ND cohort used for volumetric analysis. This cohort includes all participants processed using the fine-tuned ASCHOPLEX model following validation. Values are presented as mean ± standard deviation for age and counts for sex (female/male) *Abbreviations:* CU : cognitively unimpaired; AD : Alzheimer’s disease; PD : Parkinson’s disease.

| Diagnostic category | N | Age (mean ± SD) | Sex (F/M) |
| --- | --- | --- | --- |
| CU | 70 | 70.6 ± 6.5 | 67 / 7 |
| AD | 109 | 75.6 ± 7.8 | 46/ 63 |
| PD | 98 | 69.3 ± 7.6 | 29 / 69 |

### 2.2 MRI acquisition

T1-weighted images were acquired across 32 sites using vendor-specific COMPASS-ND protocols on Siemens, GE, and Philips scanners. Imaging was performed using harmonized 3D inversion-prepared gradient echo sequences (e.g., MPRAGE, SPGR/BRAVO, and TFE) with isotropic 1.0 mm resolution and whole-brain coverage, following consortium guidelines^25^ (https://www.cdip-pcid.ca/protocol). Although acquisition parameters varied across vendors, they were broadly comparable (e.g., TE ∼2–3 ms, TI ∼900 ms, flip angle ∼8– 12°) and employed parallel imaging (GRAPPA/ASSET/SENSE), ensuring consistent T1-weighted contrast suitable for structural segmentation.

### 2.3 Manual ChP segmentation

A representative subset of 60 COMPASS-ND T1-weighted images (balanced across diagnoses, sex and age) were independently annotated by three trained raters (MS, LJT and FD, experience: 1-2 years) following a standardised segmentation protocol (section 2.3.1) and using Display from the MINC toolkit (v1.9.18, https://bic-mni.github.io/). All raters were blinded to diagnostic categories. A joint consensus review was conducted using an established set of guidelines, with voxel-level exclusion criteria applied to ensure accurate delineation of ChP boundaries (section 2.3.1). Remaining discrepancies were resolved with a senior neuroradiologist (experience > 20 years). Final consensus masks (*N* = 60, 20 CU, 20 AD, 20 PD) served as the reference ground truth for benchmarking ASCHOPLEX segmentation performance and for model fine-tuning.

#### 2.3.1 Segmentation protocol and guidelines

Manual segmentation of the ChP was performed on T1-weighted (T1w) MRI scans to generate reference masks for model development and validation. Segmentation was restricted to the lateral ventricles, where the ChP is most clearly visualized; ChP within the third and fourth ventricles was not included. The ChP was identified as a hyperintense structure within the lateral ventricles relative to surrounding cerebrospinal fluid (CSF), which appears hypointense on T1w images. Segmentation was conducted primarily in the axial plane, with sagittal and coronal views used to confirm anatomical boundaries and ensure consistency across slices. Voxels were included if they were consistent with ChP signal characteristics and demonstrated continuity across adjacent slices or orthogonal views, with the aim of capturing the ChP as a continuous anatomical structure while avoiding over-segmentation, apart from instances when ChP was fragmented. Voxels affected by partial volume effects with CSF or adjacent tissue were excluded. Regions corresponding to the fornix were excluded unless indistinguishable from ChP based on anatomical continuity and signal characteristics. Co-registered FLAIR images were used as an adjunct in cases of low contrast or ambiguity, particularly to distinguish ChP from CSF and to identify cystic regions, which were excluded unless clearly integrated within the ChP structure. All segmentations were performed by trained raters (N=3) following these standardized criteria and were reviewed to ensure anatomical plausibility and consistency across views.

### 2.4 Automated ChP segmentation

Automated ChP segmentation was performed using ASCHOPLEX, an ensemble deep learning framework (https://github.com/FAIR-Unipd/ASCHOPLEX) combining nnU-Net and UNETR architectures, implemented via the MONAI Auto3dSeg pipeline. The model was applied “as-is” to the testing subset of *N* = 30 (10 CU, 10 AD, 10 PD) COMPASS-ND T1-weighted images for standard testing.

To assess the potential benefit of fine-tuning ASCHOPLEX to the COMPASS-ND dataset, the standard model was fine-tuned on manually annotated masks from the training and validation subset (N = 60). This fine-tuned model was then applied to the testing subset. Standard performance measures against the ground-truth manual labels were calculated for the automated segmentation outputs of both the original and fine-tuned models for statistical comparison (see Section 2.5). Following validation, the fine-tuned ASCHOPLEX model was applied to the remaining T1-w MRI scans (70 CU, 109 AD, 98 PD) to obtain automated ChP segmentations for assessing volumetric differences between diagnostic categories.

#### 2.4.1 Computational resources

Model fine-tuning and inference were conducted on a dedicated Linux workstation equipped with an NVIDIA RTX A6000 GPU (48 GB VRAM; driver version 580.95.05; CUDA 13.0) and 128 GB system memory. The ASCHOPLEX framework was implemented via MONAI Auto3dSeg using PyTorch (torch 1.13.1). The software environment included monai (v 1.0.1), nibabel (v 5.0.0), numpy (v 1.23.5), pyyaml (v 6.0), tqdm (v 4.64.1), tensorboard (v 2.11.2), fire (v 0.5.0), and einops (v 0.6.0).

### 2.5 Statistical analysis

Segmentation accuracy was quantified at the participant level before and after fine-tuning using complementary overlap- and error-based metrics. Spatial agreement between automated and manual masks was assessed using the Dice similarity coefficient and Jaccard index, which quantify the degree of voxel-wise overlap between segmentations. Precision and recall were additionally computed to characterize false-positive and false-negative behavior, respectively, providing insight into whether segmentation errors reflected over-segmentation or under-segmentation of ChP tissue. Metrics were averaged across the 30 independent test images (10 CU, 10 AD, 10 PD), and model improvement following fine-tuning was evaluated using paired-sample Wilcoxon signed-rank tests.

### 2.6 Volumetric normalization

Automated ChP volumes for the full-dataset diagnostic group comparisons were normalized to account for head size using intracranial volume (ICV). Volume distributions were tested using the Shapiro-Wilk test prior to analysis to address deviations from normality.

### 2.7 Diagnostic group comparisons

Participants diagnosed with AD with or without a vascular component were combined into a single AD group. This decision was based on preliminary analyses within the study cohort showing no significant difference in normalized ChP volume between AD and AD-vascular subgroups. The diagnostic group was therefore modeled as a three-level factor (CU, AD, PD) in all main analyses. Differences in normalized ChP volume derived from automated segmentations between CU, AD, and PD groups were assessed using a linear model: ChPnorm=β0+β1(Diagnosis)+β2(Sex)+□ where “Diagnosis” includes CU, AD and PD. Sex was treated as a fixed covariate. Post-hoc pairwise tests with Tukey-adjusted multiple comparisons were performed to identify group-specific differences.

## 3. Results

### 3.1 ASCHOPLEX segmentation performance significantly improved after fine-tuning

Table 3 presents the Dice, Jaccard, precision and recall performance measures for the standard and fine-tuned ASCHOPLEX segmentations. Application of the standard ASCHOPLEX model to 30 COMPASS-ND T1-weighted images demonstrated moderate segmentation accuracy with substantial inter-subject variability (see Table 3).

**Table 3:**
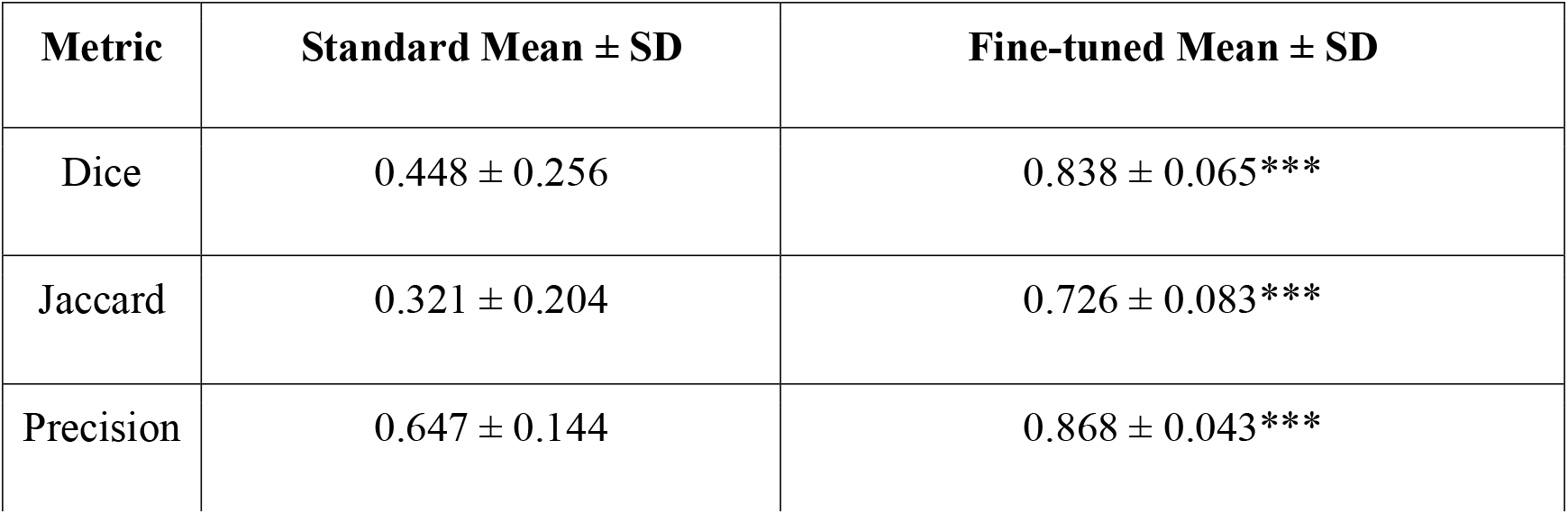
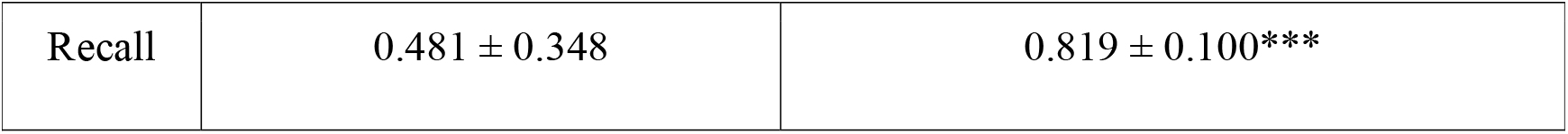
Improvement in ChP segmentation performance after ASCHOPLEX fine-tuning. Segmentation accuracy metrics (Dice, Jaccard, precision, recall; mean ± SD) are reported for standard and fine-tuned models across 30 test subjects; statistical significance was assessed using paired Wilcoxon signed-rank tests where *** indicates *p* < 0.0001.

| Metric | Standard Mean ± SD | Fine-tuned Mean ± SD |
| --- | --- | --- |
| Dice | $0.448 \pm 0.256$ | $0.838 \pm 0.065^{***}$ |
| Jaccard | $0.321 \pm 0.204$ | $0.726 \pm 0.083^{***}$ |
| Precision | $0.647 \pm 0.144$ | $0.868 \pm 0.043^{***}$ |
| Recall | $0.481 \pm 0.348$ | $0.819 \pm 0.100^{***}$ |

Following fine-tuning using 60 manually segmented consensus files, segmentation performance improved significantly (Figure 1) across all evaluated metrics (Table 3). Mean Dice improved by 0.39 and Jaccard by 0.41, indicating substantially greater spatial overlap with manual ground truth. Precision improved by 0.22, reflecting reduced false-positive segmentation, while recall increased by 0.34, indicating improved sensitivity to ChP tissue.

**Figure 1.**
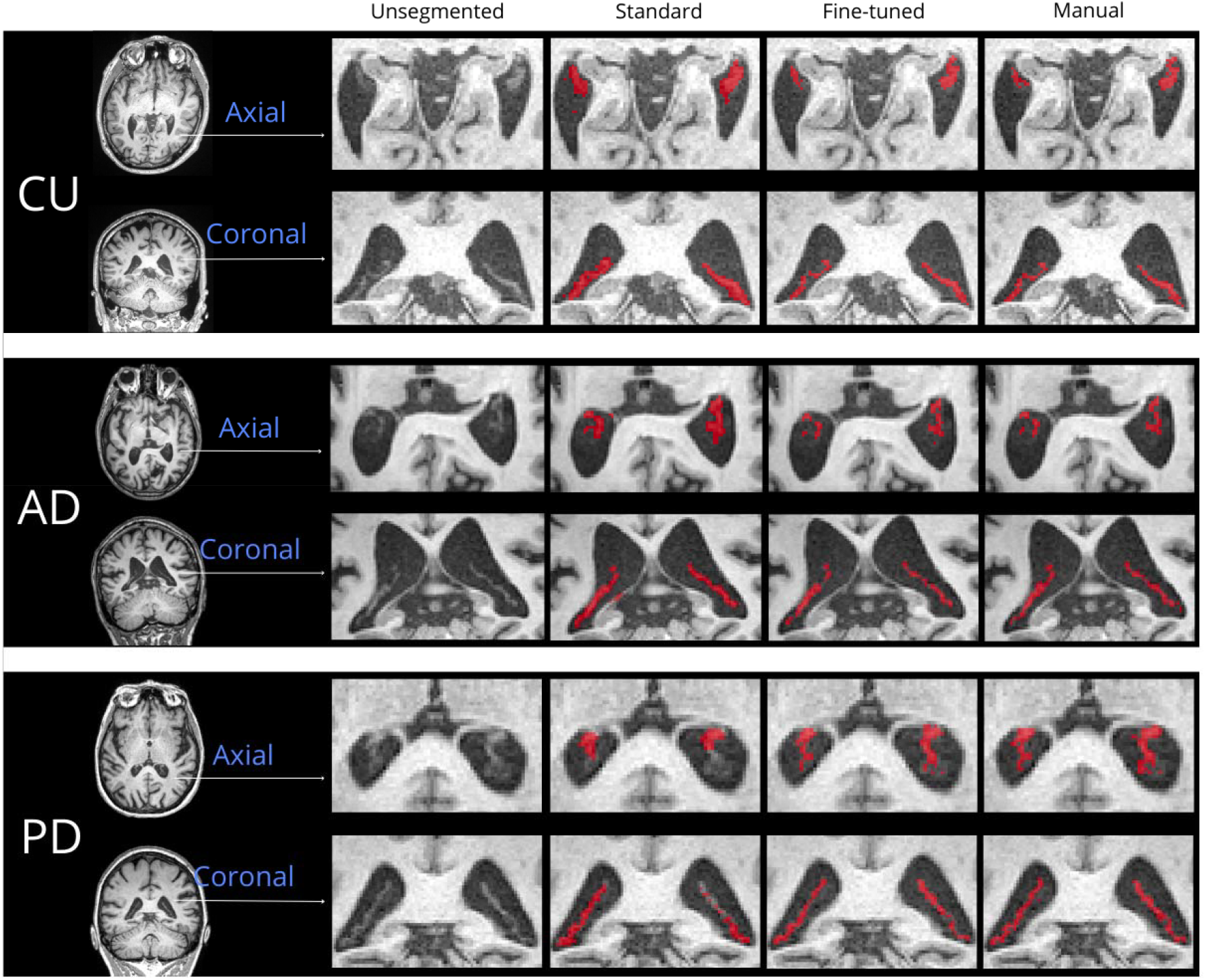
Visual comparison of ChP segmentation using standard ASCHOPLEX, fine-tuned ASCHOPLEX, and manual consensus masks. *Abbreviations:* AD: Alzheimer’s disease; ChP: choroid plexus; CU: cognitively unimpaired; PD: Parkinson’s disease.

Paired Wilcoxon signed-rank tests confirmed significant improvements after fine-tuning for Dice (*p* = 1.9 × 10^−9^), Jaccard (*p* = 1.9 × 10^−9^), precision (*p* = 6.2 × 10^−8^), and recall (*p* = 9.2 × 10^−6^), demonstrating robust and consistent gains in segmentation accuracy across subjects (Table 3).

### 3.2 Distribution and normalization of ChP volumes

Normalized ChP volumes were evaluated in the cohort comprising 70 CU, 109 AD, and 98 PD participants. Normalized ChP volumes were expressed as unitless ratios of ChP volume to ICV, scaled by 10^6^ to improve interpretability. The distribution of normalized ChP volume showed mild deviation from normality (Shapiro-Wilk test: W = 0.984, *p* = 0.0034) (Figure 2: A1, A2), with Q–Q plots indicating modest tail deviations. Log transformation did not improve normality and instead resulted in greater deviation from the theoretical distribution (W = 0.959, *p* = 4.27 × 10□□). Accordingly, all analyses were conducted using the raw normalized ChP volume values. Estimated marginal means (averaged over sex) were 1007 (95% CI: 939–1076) in CU, 1280 (95% CI: 1230–1330) in AD, and 1076 (95% CI: 1021–1130) in PD.

**Figure 2:**
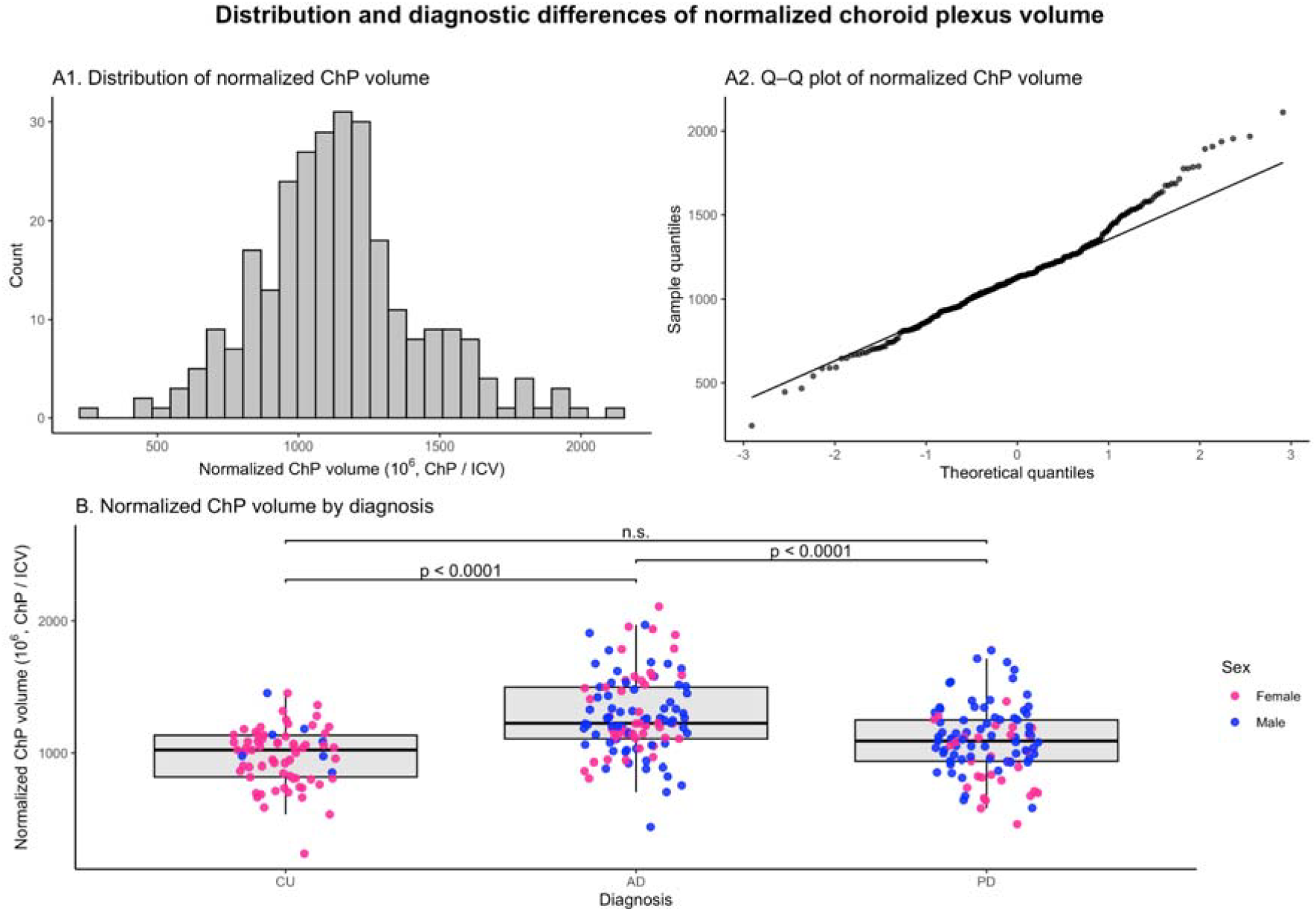
Diagnostic group differences in normalized ChP volume derived from automated ASCHOPLEX segmentations. (A1) Distribution of normalized ChP volume (ChPnorm; raw ChP volume divided by intracranial volume) stratified by sex. (A2) Q-Q plot demonstrating non-normality of ChPnorm. (B) ChPnorm across diagnostic groups (CU, AD, PD); boxplots show median and interquartile range, with individual participants overlaid and colored by sex. Brackets indicate statistically significant group differences. *Abbreviations:* AD: Alzheimer’s disease; ChP : choroid plexus; ChPnorm : normalized choroid plexus volume; CU: cognitively unimpaired; ICV: intracranial volume; PD: Parkinson’s disease; Q-Q: quantile-quantile.

### 3.3 ChP volume differs between diagnostic groups

Normalized ChP volume differed significantly across diagnostic groups (*F*(3,273) = 21.66, *p* = 1.29 × 10^−12^). Participants with AD exhibited significantly larger normalized ChP volumes compared with CU individuals (β = 272.8, *p* < 0.0001). In contrast, normalized ChP volume did not differ significantly between PD and CU participants (β = 68.5, *p* = 0.14).

Tukey-adjusted post-hoc comparisons confirmed significantly higher normalized ChP volume in AD compared with both CU (*p* < 0.0001) and PD (*p* < 0.0001) (Figure 2:B), while PD and CU did not differ significantly (*p* = 0.31).

Sex showed a modest association with normalized ChP volume, with males exhibiting slightly higher values than females (β = 64.3, *p* = 0.076), although this effect did not reach statistical significance. A diagnosis-by-sex interaction was not statistically significant, indicating that the diagnostic group differences in ChP volume (AD > PD = CU) did not differ by sex. Within-group analyses indicated a significant difference in normalized ChP volume in males and females in the PD group (*p* = 0.0005), but not in CU or AD participants.

## 4. Discussion

In this work, we provide the first systematic evaluation and adaptation of a deep learning based ChP segmentation framework across multiple neurodegenerative diagnoses in a multi-center cohort. While the standard ASCHOPLEX model demonstrated only moderate performance when applied directly to COMPASS-ND data, fine-tuning using expert manual annotations substantially improved segmentation accuracy and consistency, enabling robust ChP delineation across heterogeneous diagnoses and acquisition sites. Using volumes derived from the fine-tuned automated segmentations, we further demonstrate that normalized ChP volume is significantly larger in AD compared with CU and PD participants, consistent with prior reports of ChP enlargement in AD^1,2,7^.

### 4.1 Deep learning-based ChP segmentation in heterogeneous neurodegenerative cohorts

Recent advances in deep learning have improved neuroanatomical segmentation through data-driven modeling of spatial^22^ and contextual features^23^, with architectures such as nnU-Net^22^ enabling automated optimization of preprocessing and network design, and transformer-based models such as UNETR^23^ capturing long-range anatomical dependencies. However, segmentation of small, low-contrast structures such as the ChP remains challenging due to class imbalance, boundary ambiguity, and variability in imaging protocols. The ASCHOPLEX^19^ framework addresses these challenges by combining an ensemble of nnU-Net and UNETR architectures with optimized loss functions and a fine-tuning pipeline, enabling improved local feature extraction and global contextual modeling^19^.

When applied directly to our COMPASS-ND subset, standard ASCHOPLEX without additional training was not sufficiently reliable across test cases. This is notable because ASCHOPLEX was designed as a generalizable framework and previously outperformed commonly used approaches such as FreeSurfer and Gaussian mixture model segmentation, while also incorporating a user-accessible fine-tuning workflow to address domain shift^19^. Our findings extend prior work^19,26^ by showing that a model trained primarily on controls and multiple sclerosis data from two scanners (both Philips) benefits from dataset-specific adaptation when deployed in a multi-site neurodegenerative cohort, accounting for the inherent scanner-related variability in image characteristics across different MRI scanner brands, models, software versions and coils, despite the use of a standardized acquisition protocol.

Fine-tuning using diagnostically balanced, multi-site expert consensus annotations substantially improved segmentation accuracy and consistency across CU, AD, and PD participants. Improvements were observed across complementary overlap- and error-based metrics, indicating not only better spatial agreement but also a meaningful reduction in both over- and under-segmentation.

By reducing the reliance on labor-intensive manual segmentation while maintaining anatomical precision, the fine-tuned ASCHOPLEX framework provides a scalable and reproducible approach for ChP quantification. Such automated pipelines are particularly important for large multi-site studies such as COMPASS-ND. To facilitate reproducibility and external validation, the fine-tuned model and inference pipeline can be made available upon request (https://huggingface.co/MINDlabmtl/Aschoplex_compassnd_chp_finetuned).

### 4.2 Diagnosis-specific ChP volume alterations

Using automated volumes derived from the fine-tuned ASCHOPLEX segmentations, we observed significantly greater normalized ChP volumes in AD participants compared with both CU and PD individuals, adjusting for sex. These findings align with prior reports of ChP enlargement in AD^1,2,7^ and support the hypothesis that ChP structural changes reflect disease-specific pathophysiological processes, potentially related to impaired CSF production, inflammation, and impaired clearance of amyloid-β^2,27,28^. In contrast, ChP volume did not differ significantly between PD and CU participants. This lack of enlargement of ChP in PD aligns with recent comparative studies^1^, where ChP volume changes have been reported to be reduced^10^, increased^11,12^, or not different from controls^13,29^. These variations may reflect differences in cohort characteristics and disease stage, but may also arise from methodological variation across studies, including imaging and segmentation procedures^10–12^. Sex was also associated with ChP volume, with males exhibiting slightly larger normalized volumes than females. No diagnosis-by-sex interaction was observed, indicating that this effect was broadly consistent across groups. Although exploratory within-group comparisons suggested a difference in the PD group, this finding should be interpreted cautiously, as it is not supported by a significant interaction and may reflect differences in sample size and sex distribution across groups, particularly the low number of males in the CU group. This finding highlights the importance of considering biological covariates when analyzing ChP^7,30^.

### 4.3 Limitations

Several limitations merit consideration. First, fine-tuning and evaluation relied on a limited number of manually annotated cases (*N* = 90), although these were balanced across diagnoses. Larger annotated datasets may further improve robustness, particularly for rare or extreme morphological variants. Second, this analysis was restricted to cross-sectional T1-weighted volumetry. Future longitudinal studies are required to assess the sensitivity of serial measurements of ChP volume to longitudinal change, and to characterize ChP dynamics throughout the disease courses. Third, APOE genotype was not incorporated into the statistical model. Although APOE has been potentially associated with ChP enlargement^31^, the limited number of ε4 allele carriers in this cohort precluded a sufficiently powered analysis.

## 5. Conclusion

We successfully fine-tuned ASCHOPLEX using manually annotated masks, resulting in substantial improvements in segmentation accuracy for ChP delineation across CU, AD, and PD participants. Using the automated measurements, we show that normalized ChP volume is significantly increased in AD compared with both CU and PD participants. Our results are consistent with prior findings in the literature and further demonstrate that the refined model can be leveraged for reliable and scalable ChP quantification of T1-w MRI in heterogeneous neurodegenerative cohorts. Together, these findings highlight the value of automated ChP segmentation as a practical tool for studying structural alterations of the ChP and its potential role as an imaging marker in neurodegenerative diseases.

## Data Availability

The data used in this study were obtained from the Canadian Consortium on Neurodegeneration in Aging (CCNA) COMPASS-ND cohort and are not publicly available due to data sharing and privacy restrictions. Access to the dataset may be granted upon reasonable request and approval from the CCNA data access committee. The ASCHOPLEX framework used for automated segmentation is publicly available at: https://github.com/FAIR-Unipd/ASCHOPLEX.
The fine-tuned model weights and inference pipeline developed in this study will be made publicly available via a dedicated repository upon acceptance to a journal to facilitate reproducibility: https://huggingface.co/MINDlabmtl/Aschoplex_compassnd_chp_finetuned. In the interim, these resources can be made available upon reasonable request.

https://github.com/FAIR-Unipd/ASCHOPLEX

https://ccna-ccnv.ca/publications-process-and-data-access/

## 6. Data and Code Availability

The data used in this study were obtained from the Canadian Consortium on Neurodegeneration in Aging (CCNA) COMPASS-ND cohort and are not publicly available due to data sharing and privacy restrictions. Access to the dataset may be granted upon reasonable request and approval from the CCNA data access committee. The ASCHOPLEX framework used for automated segmentation is publicly available at: https://github.com/FAIR-Unipd/ASCHOPLEX.

The fine-tuned model weights and inference pipeline developed in this study will be made publicly available via a dedicated repository upon acceptance to a journal to facilitate reproducibility: https://huggingface.co/MINDlabmtl/Aschoplex_compassnd_chp_finetuned. In the interim, these resources can be made available upon reasonable request.

## 7. Author Contributions

AB, MS, and SN conceived the study. MS performed data processing, model implementation, and statistical analyses, contributed to manual segmentation, and led the literature review and manuscript preparation. FD led the manual segmentation efforts, contributed to data annotation, and assisted with the literature review and manuscript drafting. LJT contributed to manual segmentation and assisted with manuscript drafting. DA provided imaging methodology expertise and technical guidance. AB and SN supervised the study, contributed to study design, and critically revised the manuscript. AB secured the funding supporting the work. All authors reviewed and approved the final manuscript.

## 10. Acknowledgements

This research used data from CCNA, which was supported by a grant from the Canadian Institutes of Health Research, with funding contributions from various partner organizations.

